# Autoantibodies targeting the enteric nerve and non-myelin epitopes in relapsing-remitting multiple sclerosis: diagnostic relevance and viral mimicry

**DOI:** 10.1101/2025.06.23.25330145

**Authors:** Abbas F. Almulla, Aristo Vojdani, Muslimbek G. Normatov, Yingqian Zhang, Drozdstoj Stoyanov, Michael Maes

**Affiliations:** Sichuan Provincial Center for Mental Health, Sichuan Provincial People’s Hospital, School of Medicine, University of Electronic Science and Technology of China, Chengdu 610072, China; Key Laboratory of Psychosomatic Medicine, Chinese Academy of Medical Sciences, Chengdu 610072, China; Medical Laboratory Technology Department, College of Medical Technology, The Islamic University, Najaf, 31001, Iraq; Immunosiences Lab, Inc., Los Angeles, CA 90035, USA; Cyrex Laboratories, LLC, Phoenix, AZ 85034, USA; Department of Pathology, Faculty of Medicine, Saint Petersburg State University, 199034 Saint Petersburg, Russia; Department of Psychiatry, Faculty of Medicine, Chulalongkorn University, Bangkok, Thailand; Cognitive Fitness and Technology Research Unit, Faculty of Medicine, Chulalongkorn University, Bangkok, Thailand; Department of Psychiatry, Medical University of Plovdiv, Plovdiv, Bulgaria; Research Institute, Medical University Plovdiv, Plovdiv, Bulgaria; Kyung Hee University, 26 Kyungheedae-ro, Dongdaemun-gu, Seoul 02447, Republic of Korea

**Author notes:** **Corresponding author:** Prof. Dr. Michael Maes, M.D., Ph.D., Sichuan Provincial Center for Mental Health, Sichuan Provincial People’s Hospital, School of Medicine, University of Electronic Science and Technology of China, Chengdu 610072, China, **<u>Michael Maes Google Scholar profile,</u>** Highly cited author: 2003-2023 (ISI, Clarivate) Highly ranked scientist (lifetime and last 5 years), ScholarGPS: Worldwide #1 in molecular neuroscience; #1/4 in pathophysiology, Expert worldwide medical expertise ranking, Expertscape (December 2022), worldwide: #1 in CFS; #1 in oxidative stress; #1 in encephalomyelitis; #1 in nitrosative stress; #1 in nitrosation; #1 in tryptophan; #1 in aromatic amino acids; #1 in stress (physiological); #1 in neuroimmune; #2 in bacterial translocation; #3 in inflammation; #4-5: in depression, fatigue, and psychiatry. Joined first authorships (Abbas F. Almulla, Aristo Vojdani, Muslimbek G. Normatov).

**Keywords:** Autoimmunity, Multiple sclerosis, Enteric Nervous System, Autoantibodies, Molecular Mimicry, Herpesvirus 6, Epstein-Barr Virus

## Abstract

**Background:** Relapsing-remitting multiple sclerosis (RRMS) involves autoimmune responses against central nervous system (CNS) self-epitopes, potentially triggered by Epstein–Barr virus (EBV) and Human Herpesvirus 6 (HHV-6) reactivation.

**Objectives:** To evaluate IgG/IgA/IgM responses targeting the enteric nerve and non-myelin antigens in RRMS and investigate associations with EBV and HHV-6 markers and explore molecular mimicry between viral and host antigens.

**Methods:** The study included 55 RRMS patients and 63 matched healthy controls. ELISA is used to examine IgG/IgA/IgM levels against nine non-myelin self-epitopes and viral proteins (EBNA-1, dUTPases of EBV and HHV-6). Luminex immunoassay is used to quantify cytokines, chemokines, and growth factors. Disability was evaluated using the Expanded Disability Status Scale (EDSS) and Multiple Sclerosis Severity Score (MSSS). In-silico molecular mimicry was analyzed using UniProt, AlphaFold, PyMol, Alignmentaj, and the Immune Epitope Database (IEDB), comparing 41 viral and 36 ENS non-myelin antigens.

**Results:** IgG/IgA/IgM levels targeting the enteric nerve and non-myelin antigens were significantly higher in RRMS than in controls. A large part of the variance in the EDSS and MSSS scores (>60%) was explained by IgG-chondroitin sulfate, IgM-Asialo-ganglioside, and IgG-enteric nerve. There were highly significant correlations between autoimmunity to those self-antigens and either immune profiles indicating immune activation or Ig-responses to EBNA-1, and EBV/HHV-6 DUTPases. Molecular mimicry analysis confirmed the association between EBV/HHV-6 and the self-antigens by identifying shared pentapeptides, which might cause T-cell immunogenicity.

**Conclusion:** RRMS is characterized by autoimmune responses targeting the enteric nerve and non-myelin proteins. EBV and HHV-6 reactivation contribute via molecular mimicry mechanisms.

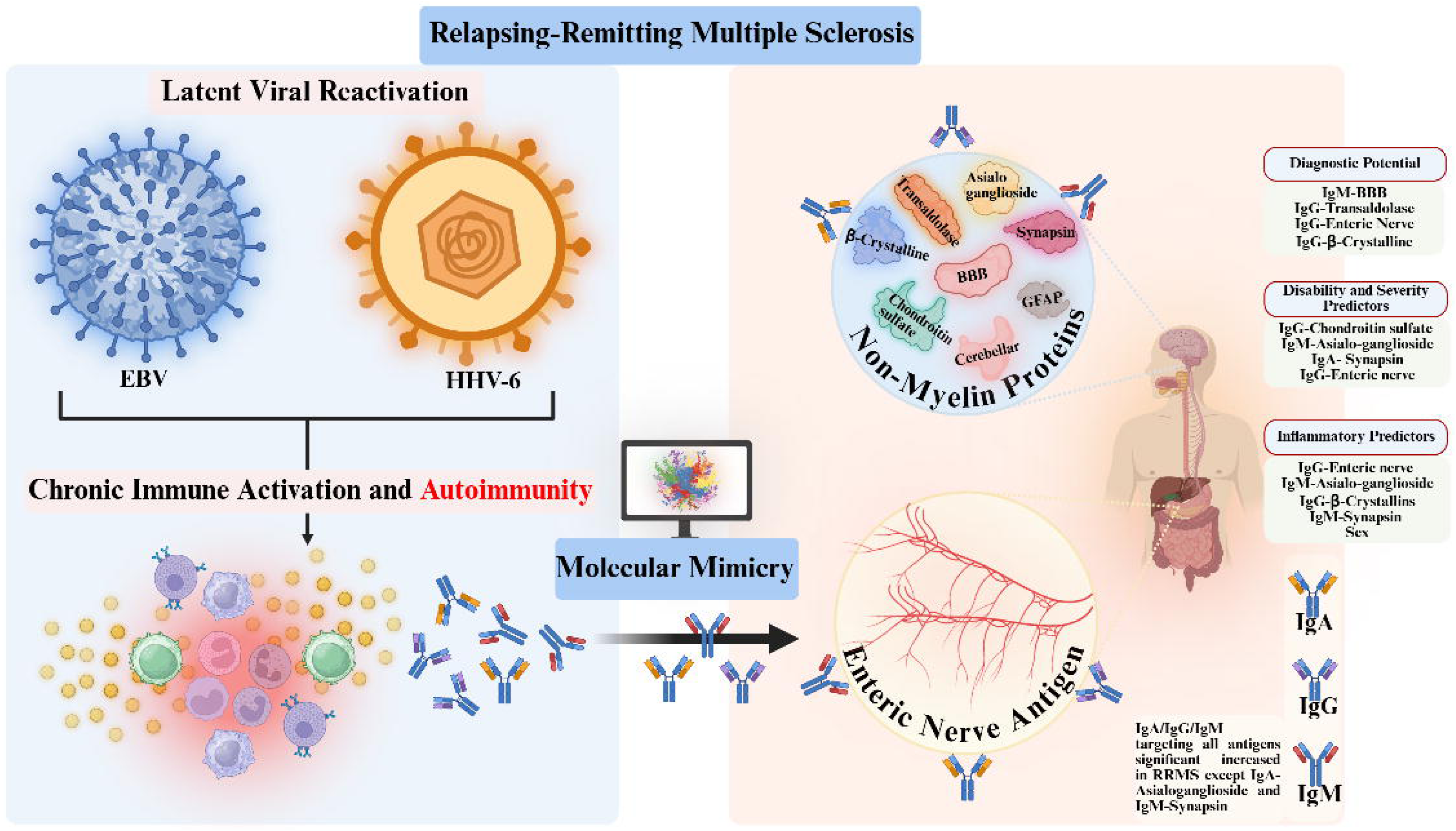

## Introduction

Relapsing-remitting multiple sclerosis (RRMS) is the predominant subtype of multiple sclerosis (MS), impacting around 85% of individuals with MS [1, 2]. It is defined by episodes of neurological impairment that are succeeded by partial or complete remission [3]. The global burden of MS has risen to 1.89 million individuals, indicating its significant impact as a neurological and socioeconomic issue [4, 5]. Some studies indicate a correlation between relapse frequency and increased disability [6].

Autoimmune responses targeting CNS proteins are regarded as fundamental to the pathophysiology of RRMS [7–10]. Recent findings indicate markedly increased levels of IgG, IgA, and IgM antibodies targeting myelin basic protein (MBP), myelin-associated glycoprotein, and myelin oligodendrocyte glycoprotein (MOG) [7]. However, autoimmunity in MS likely extends beyond classical myelin self-epitopes.

Autoantibodies directed against enteric glial and neuronal structures have been identified in experimental models as well as in patient sera [11–13]. Additionally, Banati revealed that at least one antibody that targets common GI mucosal antigens is present in 28% of MS patients [14]. Histopathological findings indicate that gliosis and neuronal damage in the ENS may occur prior to CNS demyelination in certain patients [11]. The findings suggest that the enteric nervous system (ENS) may function as an early site of autoimmune activity in MS.

Patients with MS exhibit antibodies against gangliosides, especially anti-GD2-like IgM, which are linked to increased disease severity [15]. Moreover, antibodies targeting αB-crystallin (found primarily in the lens of the eye) were detected in the cerebrospinal fluid (CSF) of MS patients and in sera from animal models of MS [16]. Serum and CSF of MS patients showed high-affinity autoantibodies against transaldolase, which is highly expressed in oligodendrocytes and plays a key role in the pentose phosphate pathway (PPP) [17–19]. BBB dysfunction in MS allows immune cell infiltration alongside lesion formation and disease progression [20, 21]. Autoantibodies targeting BBB components, including galectin-3 and GRP78, have been identified in MS and other neuroimmunological diseases [22]. Additional non-myelin-associated autoantibodies were underexplored in RRMS including those directed against the BBB proteins (occludin and claudin), and chondroitin sulfate, glial fibrillary acidic protein (GFAP), and cerebellar protein 2. Nonetheless, the autoimmunity directed against peripheral components, especially those associated with the ENS, gangliosides, and β-crystallin have not been thoroughly explored.

Chronic viral infections, particularly Epstein-Barr virus (EBV) and human herpesvirus 6 (HHV-6), are significantly associated with the onset and progression of MS [23–25]. Patients with RRMS often show increased levels of antiviral antibodies, suggesting viral reactivation associated with relapse episodes and clinical decline [25, 26]. HHV-6 active infection is more common during RRMS relapses than remissions, and is associated with earlier disease onset and greater disability progression [27, 28].

Reactivation of EBV and HHV-6 can be associated with activation of M1 macrophages and T helper (Th)17 cells, both of which are involved in the pathogenesis of MS [25, 26, 29]. IgA responses to GlialCAM were found to correlate with antibodies to EBNA and HHV-6 dUTPase, indicating the potential role of molecular mimicry [7, 30]. Recently, in-silico findings identified common pentapeptides between EBV/HHV-6 viral antigens and CNS proteins, providing mechanistic insights into the activation of autoreactive T and B cells [31].

This study aims to investigate whether serum levels of IgA, IgM, and IgG autoantibodies targeting the enteric nerve, Asialo-gangliosides, transaldolase, β-crystallins, and chondroitin sulphate, the blood-brain barrier (BBB), cerebellar, synapsin, glial fibrillary acidic protein (GFAP) are higher in RRMS patients than in healthy controls. Additionally, we examine whether these autoantibodies are associated with clinical disabilities, immune activation (as assessed with serum profiles of cytokines, chemokines, and growth factors), and signs of EBV and HHV-6 reactivation (as assessed with EBNA, and EBV/HHV-6 dUTPAses). Finally, an in-silico analysis is performed to investigate possible molecular mimicry between ENS peptides and EBV/HHV-6 proteins.

## Subjects and Methods

### Subjects

Fifty-five patients with RRMS were recruited for this case-control intervention at the Neuroscience Centre in Al-Sader Medical City, Al-Najaf, Iraq. A senior neurologist recognized for his expertise in MS confirmed the diagnosis of RRMS. We adhered to the new McDonald criteria [32]. To maintain demographic comparability, 63 neurologically healthy individuals were recruited from the same location for the control group. These subjects were hospital staff, their relatives, and acquaintances of patients with RRMS.

The subsequent neuro-psychiatric conditions were explicitly excluded from inclusion: major depressive disorder, bipolar disorder, generalized anxiety disorder, obsessive-compulsive disorder, panic disorder, schizophrenia, post-traumatic stress disorder, non-multiple sclerosis, delirium, neurodegenerative disorders such as Parkinson’s disease and Alzheimer’s disease, substance use disorders (excluding nicotine dependence), chronic fatigue syndrome/myalgic encephalomyelitis, and autism spectrum disorders. We excluded major medical illnesses such as type 1 diabetes mellitus, rheumatoid arthritis, inflammatory bowel disease, systemic lupus erythematosus, psoriasis, cancer, thyroid dysfunctions, renal or hepatic disease, and clinically significant pulmonary or cardiovascular conditions.

Before engaging in the study, all participants (or their legal guardians, when applicable) provided written informed consent. The Institutional Ethics Committee of the College of Medical Technology of the Islamic University of Najaf granted approval, reference number 11/2021. The study adhered to all relevant national and international standards concerning research ethics, including the ICH-GCP principles, CIOMS guidelines, and the Declaration of Helsinki established by the World Medical Association. Adherence to these regulatory frameworks was accomplished to protect the rights and welfare of all human participants through supervision by the Institutional Review Board.

### Clinical Assessments

All subjects received semi-structured interviews conducted by a senior neurologist specializing in MS to get detailed clinical and demographic data. The Expanded Disability Status Scale (EDSS), a widely recognized tool developed by Kurtzke in 1983, was employed to quantify neurological disability and evaluate functional limits associated with MS [33]. The Multiple Sclerosis Severity Score (MSSS) was utilized concurrently to contextualize disability in relation to disease duration, facilitating a dynamic evaluation of disease progression; this instrument was initially introduced by Roxburgh and associates in 2005 [34]. The body mass index (BMI) of each participant was calculated by dividing weight (kg) by height (m²).

### Biomarkers assays

Blood samples were obtained from participants in a fasting condition using venipuncture, conducted between 7:30 and 9:00 a.m. utilizing sterile, disposable syringe. Following a 15-minutes incubation at room temperature for blood clotting, samples were centrifuged at 3500 revolutions per minute for 10 minutes to isolate the serum. The extracted serum was divided into Eppendorf tubes and preserved for future ELISA and Luminex assays. The enteric nerve proteins were obtained from Sigma-Aldrich (St. Louis, MO, USA), while the Asialo-ganglioside, transaldolase, β-Crystalline, the BBB proteins from Bio-Synthesis® (Lewisville, TX, USA). The former company provided synapsin, GAFP and HHV-6 dUTPase, EBV dUTPase, and the EBNA-366–406 epitope. Chondroitin sulfate and cerebellar protein 2 were purchased from CUSABIO (Houston, TX, USA). These antigens were employed as coating agents in enzyme-linked immunosorbent assays (ELISA) to assess humoral immune responses.

In the ELISA techniques, each well of a 96-well plate was coated with 100 µL of antigen solution, calibrated to an optimal concentration of 5–10 µg/mL in 0.1 M carbonate buffer at pH 9.5. After an incubation period, plates were meticulously washed and subsequently blocked with 2% bovine serum albumin (BSA) to reduce nonspecific binding. Serum samples from both RRMS patients and healthy controls were subsequently applied in duplicate—diluted 1:50 for IgA detection and 1:100 for IgG and IgM measurements—and incubated at 25°C for one hour. Following a further washing phase, enzyme-conjugated secondary antibodies were introduced, succeeded by the application of the substrate. Colorimetric alterations were quantified spectrophotometrically, and immunoreactivity indices were computed utilizing standard reference sera obtained from clinically characterized MS patients. Comprehensive methodological details have been documented in prior publications [35, 36].

Furthermore, we quantified serum levels of cytokines, chemokines, and growth factors to characterize immune-inflammatory activity, focusing on the immune-inflammatory response system (IRS) and the compensatory immunoregulatory system (CIRS). The measurements were conducted utilizing the Bio-Plex Pro™ Human Chemokine Assay platform (Bio-Rad Laboratories, Hercules, CA, USA), adhering to protocols established in prior studies [37]. ESF1, Table 1 provides a summary of the immune mediators included in the panel.

**Table 1.** Results of general linear models (GLM) which show the associations between autoimmune biomarkers and relapsing-remitting multiple sclerosis (RRMS) versus healthy controls (HC). The associations were adjusted for age, sex, BMI, and smoking.

| **Variables** | **Healthy Controls**<br>**(n=63)** | **RRMS Patients**<br>**(n=55)** | **F** | **df** | **p-value** |
| --- | --- | --- | --- | --- | --- |
| IgG-Enteric nerve | 0.792 ± 0.032 | 1.405 ± 0.034 | 165.33 | 1/112 | <0.001 |
| IgM-Enteric nerve | 0.548 ± 0.049 | 1.117 ± 0.053 | 59.12 | 1/112 | <0.001 |
| IgA-Enteric nerve | 0.539 ± 0.032 | 0.927 ± 0.035 | 62.67 | 1/112 | <0.001 |
| IgG-Asialo-ganglioside | 0.383 ± 0.097 | 1.353 ± 0.105 | 43.79 | 1/112 | <0.001 |
| IgM-Asialo-ganglioside | 0.283 ± 0.037 | 0.768 ± 0.040 | 75.09 | 1/112 | <0.001 |
| IgA-Asialo-ganglioside | 2.188 ± 12.039 | 16.578 ± 12.931 | 0.63 | 1/112 | 0.429 |
| IgG- Transaldolase | 0.262 ± 0.034 | 0.734 ± 0.037 | 84.15 | 1/112 | <0.001 |
| IgM- Transaldolase | 0.335 ± 0.040 | 0.864 ± 0.043 | 75.74 | 1/112 | <0.001 |
| IgA- Transaldolase | 0.281 ± 0.035 | 0.759 ± 0.038 | 79.97 | 1/112 | <0.001 |
| IgG- β-Crystalline | 0.448 ± 0.044 | 1.160 ± 0.047 | 115.32 | 1/112 | <0.001 |
| IgM- β-Crystalline | 0.456 ± 0.042 | 0.966 ± 0.045 | 65.58 | 1/112 | <0.001 |
| IgA- β-Crystalline | 0.484 ± 0.040 | 0.965 ± 0.043 | 62.21 | 1/112 | <0.001 |
| IgG-BBB | 0.374 ± 0.038 | 0.731 ± 0.041 | 38.94 | 1/112 | <0.001 |
| IgM-BBB | 0.370 ± 0.041 | 0.913 ± 0.044 | 76.03 | 1/112 | <0.001 |
| IgA-BBB | 0.310 ± 0.023 | 0.528 ± 0.025 | 40.11 | 1/112 | <0.001 |
| IgG- Chondroitin sulfate | 0.360 ± 0.032 | 0.868 ± 0.035 | 109.68 | 1/112 | <0.001 |
| IgM- Chondroitin sulfate | 0.350 ± 0.033 | 0.750 ± 0.036 | 64.41 | 1/112 | <0.001 |
| IgA- Chondroitin sulfate | 0.305 ± 0.029 | 0.593 ± 0.031 | 44.04 | 1/112 | <0.001 |
| IgG-Cerebellar protein 2 | 0.352 ± 0.098 | 1.269 ± 0.105 | 38.89 | 1/112 | <0.001 |
| IgM- Cerebellar protein 2 | 0.353 ± 0.040 | 0.793 ± 0.043 | 52.91 | 1/112 | <0.001 |
| IgA- Cerebellar protein 2 | $0.316 \pm 0.032$ | $0.577 \pm 0.035$ | 29.02 | 1/112 | <0.001 |
| IgG- Synapsin | $0.502 \pm 0.068$ | $0.849 \pm 0.074$ | 11.32 | 1/112 | 0.001 |
| IgM- Synapsin | $0.395 \pm 11.978$ | $18.672 \pm 12.866$ | 1.03 | 1/112 | 0.313 |
| IgA- Synapsin | $0.318 \pm 0.030$ | $0.559 \pm 0.032$ | 28.03 | 1/112 | <0.001 |
| IgG-GFAP | $0.759 \pm 0.063$ | $1.401 \pm 0.068$ | 45.26 | 1/112 | <0.001 |
| IgM-GFAP | $0.635 \pm 0.048$ | $0.974 \pm 0.052$ | 21.75 | 1/112 | <0.001 |
| IgA-GFAP | $0.557 \pm 0.055$ | $0.941 \pm 0.059$ | 21.69 | 1/112 | <0.001 |

Ig: Immunoglobulin, BBB: Blood-brain barrier, GFAP: Glial fibrillary acidic protein

Raw immunofluorescence intensities were adjusted for background signal and standardized into z-scores, following established analytical frameworks [38, 39]. The values were utilized to calculate composite indices that represent specific immune profiles [38, 40–42]. The definitions and composition of each immune profile are detailed in ESF1, Table 2.

**Table 2.**
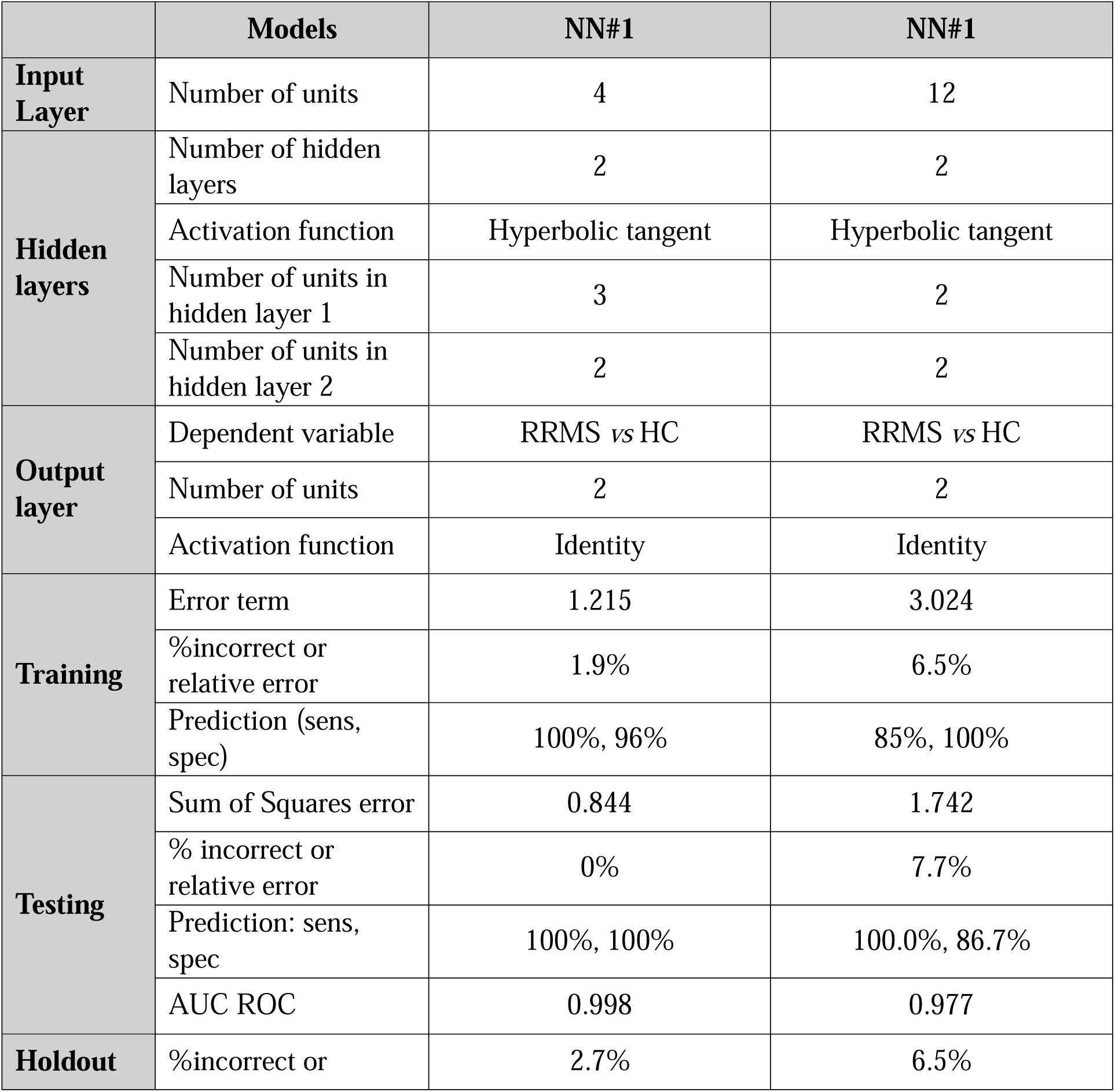
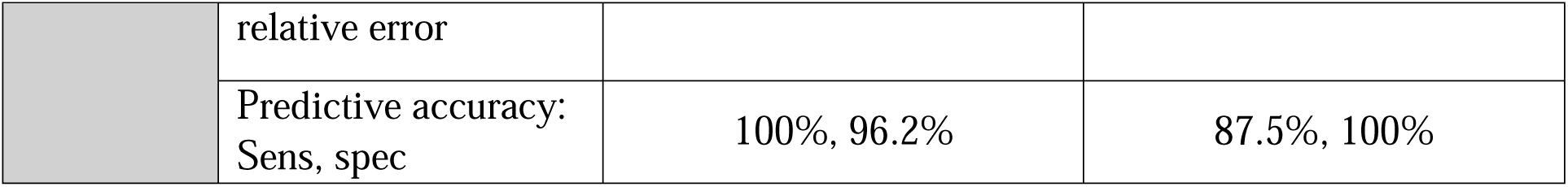
Results of neural networks (NN) with the diagnosis relapsing-remitting multiple sclerosis (RRMS) diagnoses as output variables and autoimmune biomarkers as input data.

## Data analysis

Statistical analyses were performed utilizing IBM SPSS Statistics (version 29). A preliminary sample size estimation was performed using G*Power (version 3.1.9.7), indicating that at least 73 participants are necessary to identify a significant departure of R² from zero in a fixed linear multiple regression model. This estimation was predicted on an effect size of 0.176, a power of 0.80, an alpha level of 0.05, and the incorporation of four covariates.

Group-level comparisons were examined using one-way analysis of variance (ANOVA) for continuous variables and contingency table analyses for categorical variables. The false discovery rate (FDR) p-correction was applied to mitigate type I errors in multiple comparisons. Pearson’s correlation coefficient was utilized to evaluate the correlations between continuous variables. Multivariate linear regression analysis was conducted to examine the predictive influence of autoimmune reactions to ENS antigens on clinical outcomes, encompassing disability and symptom severity indices. The model was constructed using manual entry and automatic stepwise selection techniques, using p-value thresholds of < 0.05 for variable inclusion and ≥ 0.06 for exclusion. Statistical control was used by allowing for the effects of confounders including age, sex, body mass index (BMI), and smoking status. Regression outputs comprised standardized beta coefficients, p-values, R², F-statistics, and degrees of freedom. Heteroskedasticity was tested using the White and modified Breusch–Pagan tests, while multicollinearity was evaluated through tolerance values and variance inflation factors (VIF).

Principal component analysis (PCA) was employed to diminish dimensionality specifically clinical severity ratings, disability scores, and viral antigen measurements. One principal component was extracted when the Kaiser-Meyer-Olkin (KMO) index surpassed 0.6, the first component accounted for over 50% of the total variance, and all factor loadings on the component exceeded 0.7. The principal component scores were consequently used in other statistical tests.

Multilayer perceptron neural network models were created to differentiate RRMS patients from healthy controls by examining immunoglobulin (IgG, IgA, IgM) responses to ENS and CNS autoantigens, including enteric nerve, BBB, transaldolase, β-crystallins, chondroitin sulfate and GFAP. The models employed a feedforward architecture featuring two hidden layers, each containing up to eight units, and were trained in batch mode for a maximum of 250 epochs. Training was considered complete when no more decreases in error were detected. The evaluation of model performance encompassed classification accuracy, misclassification rates, error metrics, and relative error assessments. Variable importance plots were employed to demonstrate the relative significance of each input variable.

## In Silico part

### Acquisition of Protein Structure and Mapping of Mimicry

To investigate possible molecular mimicry between EBV and HHV-6 antigens and ENS autoantigens, 41 viral proteins were chosen for structural comparison, including 28 from HHV-6 and 13 from EBV (as detailed in ESF2, Table 1 and 2). The viral antigens were methodically analyzed for common sequence motifs, particularly pentapeptides, in conjunction with 37 ENS-associated human autoantigens documented in ESF2, Table 1. The full amino acid sequences for both viral and human ENS proteins were obtained from the UniProt database [43] in FASTA format, with the relevant UniProt accession codes listed in ESF2, Table 1.

Three-dimensional models of all ENS autoantigens were obtained in PDB format from the AlphaFold Protein Structure Database [44] and subsequently visualised using PyMOL (https://www.pymol.org/) [45] to delineate the spatial distribution of identified mimicry pentapeptides on their molecular surfaces. The AlphaFold database identifiers for the ENS autoantigen structures are enumerated in ESF2, Table 2.

### Identification of Mimetic Pentapeptides

The custom software tool “Alignmentaj” (Certificate #2,023,617,186, Russian Federal Agency for Intellectual Property; source code accessible at: https://github.com/muslimb/MyProekt1, registry: https://www.elibrary.ru/item.asp?id=52295110) was utilized to systematically identify overlapping sequence segments. This program accepts amino acid sequences of viral and human proteins in FASTA format and extracts overlapping pentapeptides from the viral proteins (e.g., MSDEG, SDEGP, DEGPG). The pentapeptides are subsequently matched with the human auto-antigen sequences to pinpoint identical portions. The output enumerates common pentapeptides that represent potential molecular mimicry motifs between viral and ENS proteins (ESF1, Table 1 and 2).

### Assessment of Immunogenic Mimicry: Significance of T and B Cells

We conducted epitope analysis utilizing the Immune Epitope Database (IEDB) [46] to evaluate the potential immunogenicity of these conserved pentapeptides in provoking human immune responses. This platform consolidates empirical data on experimentally confirmed epitopes identified by human T and B lymphocytes. Pentapeptides common to herpesvirus antigens (EBV and HHV-6) and ENS proteins were analyzed to assess their potential role as immunogenic epitopes in T and B cell activation. The objective was to uncover molecular signals that may play a role in eliciting autoimmune responses via antigenic mimicry.

## Results

### Sociodemographic and clinical characteristics of participants

Table 3 in the ESF1 provides a comparative analysis of sociodemographic characteristics and disability and clinical severity indices, namely the EDSS and the MSSS, for patients with RRMS and healthy subjects. The two groups showed no significant differences in age, gender distribution, marital status, and BMI. Differences in employment and smoking status between the RRMS cohort and the control group.

**Table 3.** Results of multiple regression analysis performed in the total study group and in the patients’ group with severity rating scale scores or immune profiles as dependent variables and autoimmune responses to various antigens as explanatory variables.

| Dependent Variables | Explanatory Variables | Coefficient statistics |  |  | Model statistics |  |  |  |
| --- | --- | --- | --- | --- | --- | --- | --- | --- |
| | | $\beta$ | t | p | R<sup>2</sup> | F | df | p |
| #1 EDSS | **Model** |  |  |  | 0.719 | 146.80 | 2/115 | <0.001 |
|  | IgG-Chondroitin sulfate | 0.550 | 7.419 | <0.001 |  |  |  |  |
|  | IgM-Asialo-ganglioside | 0.354 | 4.781 | <0.001 |  |  |  |  |
| #2 MSSS | **Model** |  |  |  | 0.627 | 96.74 | 2/115 | <0.001 |
|  | IgG-Enteric nerve | 0.471 | 5.886 | <0.001 |  |  |  |  |
|  | IgM-Asialo-ganglioside | 0.387 | 4.840 | <0.001 |  |  |  |  |
| #3 EDSS (In patients only) | **Model** |  |  |  | 0.073 | 4.18 | 1/53 | 0.046 |
|  | IgA- Synapsin | 0.270 | 2.045 | 0.046 |  |  |  |  |
| #4 MSSS (In patients only) | **Model** |  |  |  | 0.219 | 14.81 | 1/53 | <0.001 |
|  | IgA-Synapsin | 0.467 | 3.850 | <0.001 |  |  |  |  |
| #5 IRS/CIRS | **Model** |  |  |  | 0.487 | 50.86 | 2/107 | <0.001 |
|  | IgG-Enteric nerve | 0.447 | 4.763 | <0.001 |  |  |  |  |
|  | IgM-Asialo-ganglioside | 0.313 | 3.340 | 0.001 |  |  |  |  |
| # 6 IRS | **Model** |  |  |  | 0.262 | 38.29 | 1/108 | <0.001 |
| | IgG-β-Crystallins | 0.512 | 6.188 | <0.001 | | | | |
| #7 CIRS | **Model** |  |  |  | 0.613 | 55.87 | 3/106 | <0.001 |
|  | IgG-Enteric nerve | 0.548 | 7.564 | <0.001 |  |  |  |  |
|  | IgM-Synapsin | 0.319 | 4.397 | <0.001 |  |  |  |  |
|  | Sex | 0.134 | 2.222 | 0.028 |  |  |  |  |

EDSS: Expanded Disability Status Scale, MSSS: Multiple Sclerosis Severity Score, Ig: Immunoglobulin, IRS: Immune response system, CIRS: Compensatory-immune-regulatory system, GFAP: Glial fibrillary acidic protein.

Alongside clinical metrics, principal component scores reflecting humoral immune responses were analyzed. In the RRMS group, PC scores for IgA, IgM, and IgG antibodies targeting EBNA, EBV dUTPase, and HHV-6 dUTPase antigens were significantly higher compared to the control group. The composite scores, designated as PC-3EBNAs, PC-3dUTPases-EBV, and PC-3dUTPases-HHV-6, were derived by extracting the first principal component from the three immunoglobulin isotypes (IgA, IgM, IgG) that target each viral antigen. Each PC score serves as a comprehensive index of immune reactivity across different antibody classes targeting a specific viral protein [7].

### Comparative analysis of ENS-targeted autoimmune profiles across study cohorts

Table 1 indicates that individuals with RRMS exhibited significantly elevated immunoglobulin responses (IgG, IgA, and IgM) to all ENS and the non-myelin self-epitopes (except two), including enteric nerve, Asialo-ganglioside, transaldolase, β-crystallin, BBB antigens, chondroitin sulphate, cerebellar proteins, synapsin, and GAFP. Notably, two antibody-antigen interactions—IgA directed against Asialo-ganglioside and IgM directed against synapsin—showed no significant differences between RRMS patients and healthy controls. The effect size analysis, employing Partial Eta Squared (η²), revealed the most significant group differences in the following order: IgG reactivity to enteric nerve proteins (η² = 0.596), IgG to β-crystallin (η² = 0.507), IgG to chondroitin sulphate (η² = 0.495), IgG to transaldolase (η² = 0.429), IgA to transaldolase (η² = 0.417), and IgM to blood-brain barrier components (η² = 0.404).

### Accuracy of the autoantibodies for RRMS

A binary logistic regression analysis was conducted to identify the primary immunological predictors of RRMS, with diagnostic status (RRMS vs. control) as the dependent variable and the control group serving as the reference category. IgG antibodies directed against enteric nerve proteins emerged as the most effective discriminative signal among all predictors. This model exhibited perfect classification performance, attaining 100% accuracy, with both sensitivity and specificity at 100%, and an effect size of 1.

In addition to the logistic regression findings, a neural network analysis in **Table 2** was performed to evaluate the predictive importance of chosen immunological markers. The initial neural architecture (NN#1) developed to differentiate RRMS patients from healthy individuals included two hidden layers, with two nodes in the first layer, three in the second, and a dual-node output layer. The error rate in the testing set was significantly lower than that in the training set, and performance indicators were consistent across the training, testing, and holdout subsets, suggesting model stability and the absence of overfitting. The model attained an accuracy of 97.3% on the holdout dataset (sensitivity = 100%, specificity = 96.2%) and a ROC area of 0.998. The variable importance plot (**Figure 1**) indicates that the key predictors were IgM-BBB, IgG-Transaldolase, and IgG-Enteric nerve.

**Figure 1.**
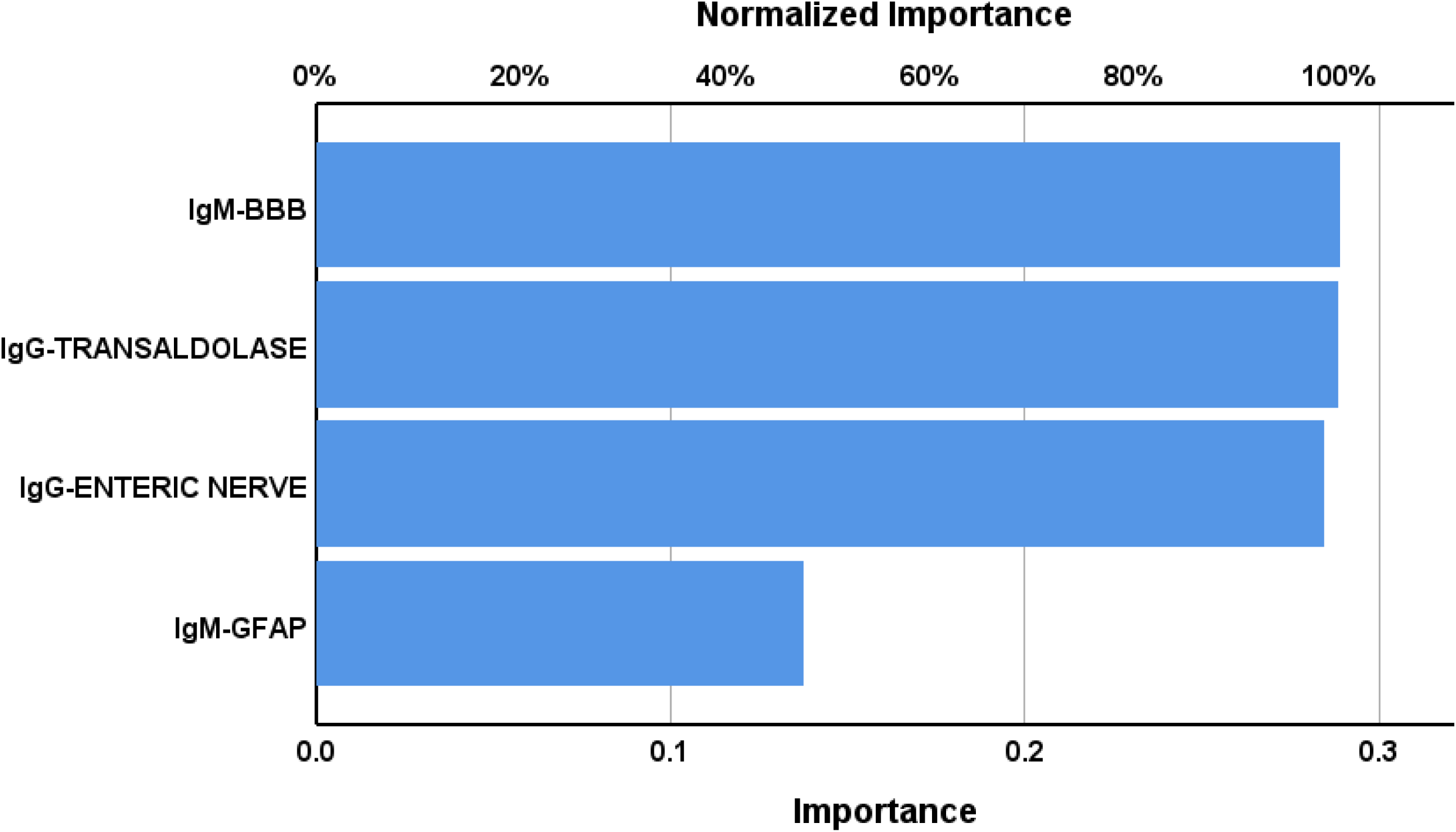
Results of neural network analysis showing the importance of the selected biomarkers. Output variables: diagnosis of relapsing remitting multiple sclerosis and healthy controls. Input variables: Immunoglobulin (Ig) responses targeting self-antigens. BBB: blood-brain barrier; GFAP: glial fibrillary acidic protein.

A second neural network (NN#2) was developed to examine the accuracy of a model that includes all antigens assayed in this study without those incorporated in model #1. This model retained a comparable architecture featuring two hidden layers, each consisting of two units, and maintained two units in the output layer as shown in Table 2. The error distribution exhibited reduced values during the testing phase compared to the training phase, with uniform error rates noted across all subsets. NN#2 attained a classification accuracy of 95.5% in the holdout sample, with a sensitivity of 87.5%, specificity of 100%, and a ROC area under the curve of 0.977. **Figure 2** demonstrates that the primary predictors in NN#2 were IgG antibodies targeting β-crystallin, succeeded by IgG responses to chondroitin sulphate.

**Figure 2.**
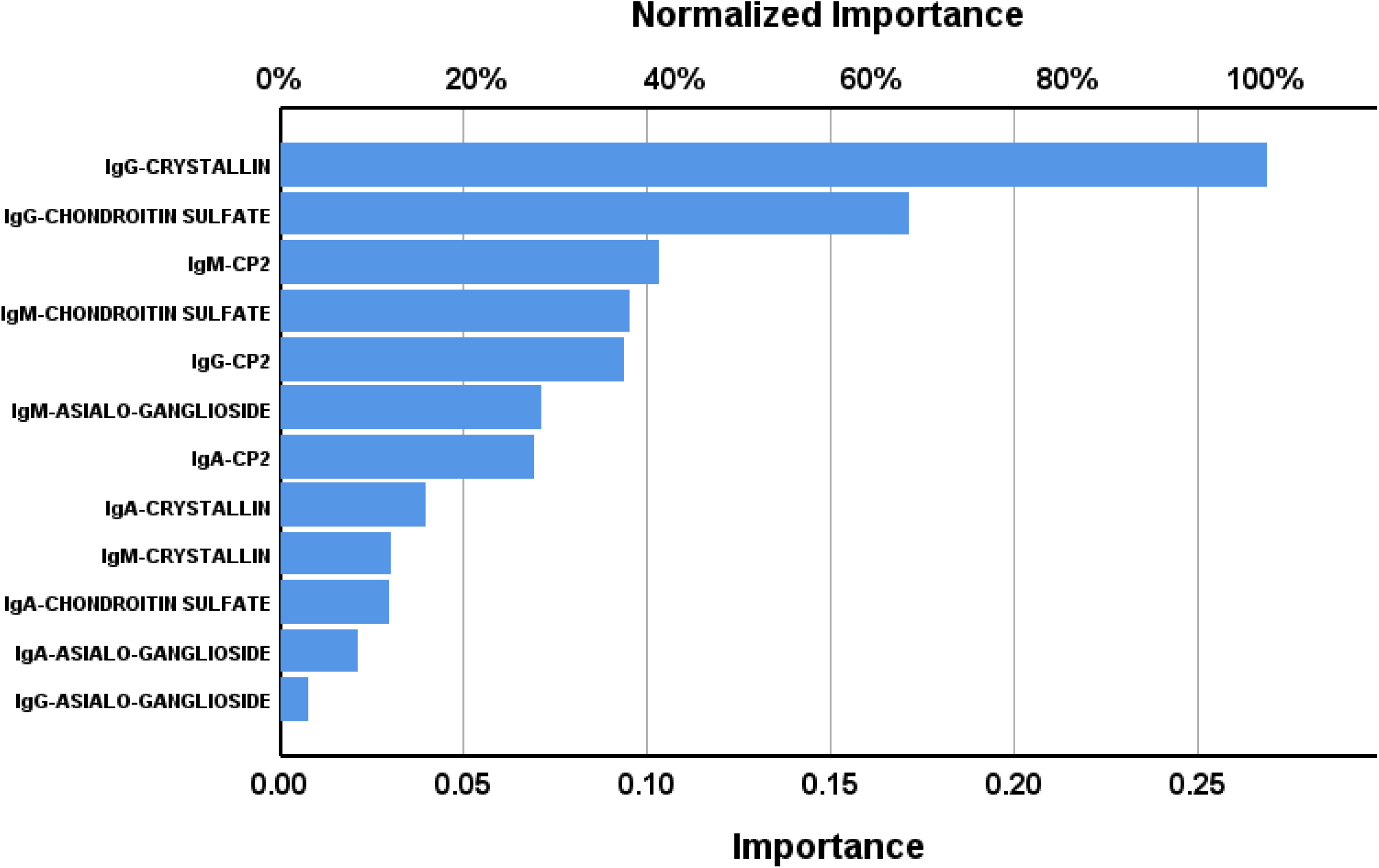
Results of neural network analysis showing the importance of the selected biomarkers. Output variables: diagnosis of relapsing remitting multiple sclerosis and healthy controls. Input variables: Immunoglobulin (Ig) responses targeting self-antigens. CP2: cerebellar protein 2.

### Prediction of RRMS severity by autoimmune responses

**Table 3** presents the results of multivariate regression models that investigate the relationships between immune-inflammatory biomarkers and clinical parameters in patients with RRMS. The dependent variables assessed in the models comprised the EDSS, MSSS, IRS/CIRS ratio, and CIRS. The explanatory data were the IGG/IgM/IgA responses to the neuronal antigens, whilst we allowed for the effects of age, sex, BMI, and smoking.

The first regression demonstrated a positive correlation between EDSS scores and both IgG-chondroitin sulphate and IgM-Asialo-ganglioside, accounting for 71.9% of the variance. **Figure 3** illustrates the partial regression plot of EDSS on IgG-chondroitin sulphate, with age and sex controlled for. In regression #2, MSSS scores were predicted by IgG-enteric nerve and IgM-Asialo-ganglioside, both of which exhibited a positive association, together explaining 62.7% of the variance. **Figure 4** presents the partial regression plot illustrating the adjusted relationship between MSSS and IgG-enteric nerve. In RRMS patients, IgA-synapsin contributed 7.3% to the variance in EDSS (regression #3), whereas regression #4 showed that this marker accounted for 21.9% of the variance in MSSS.

**Figure 3.**
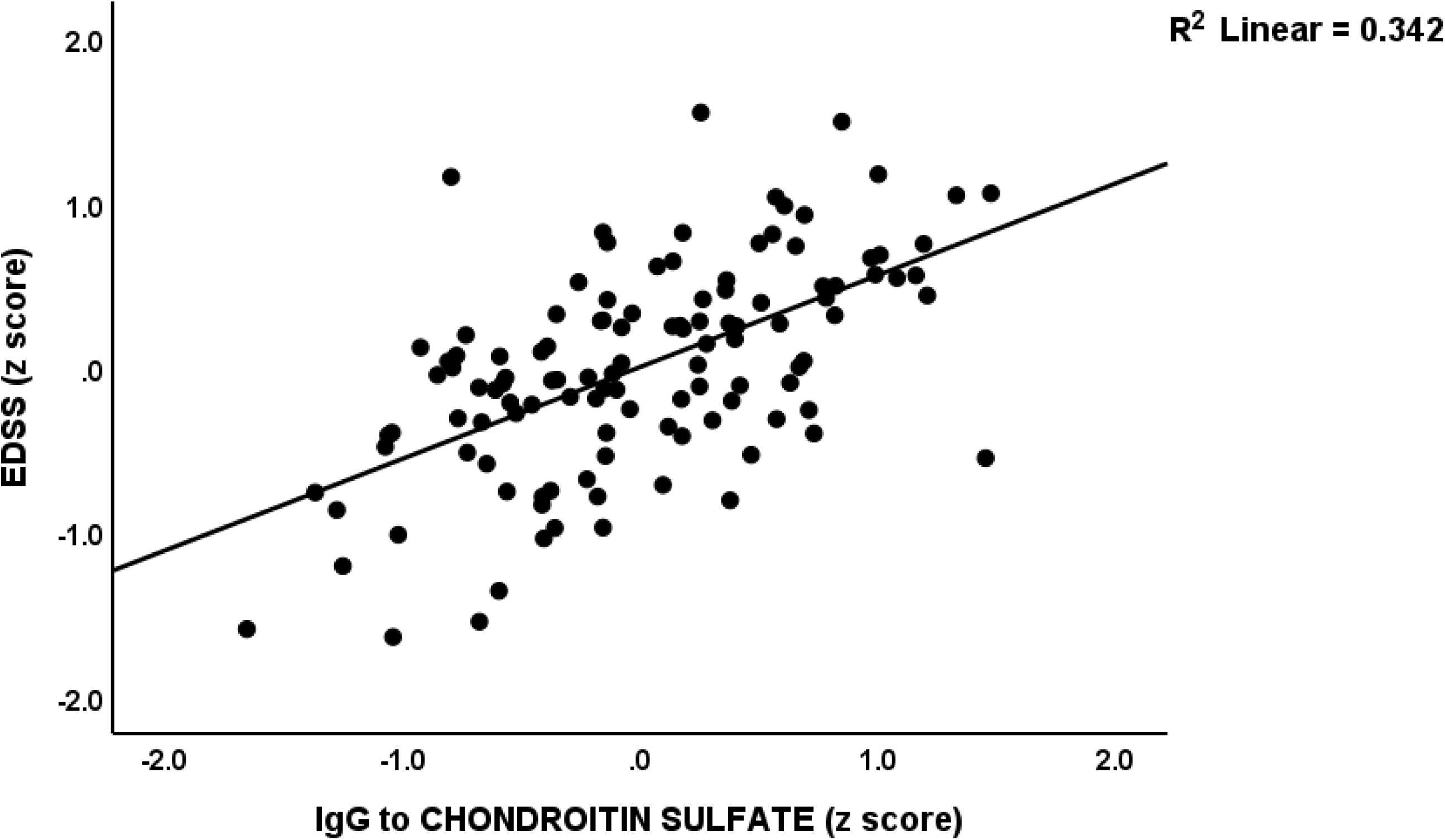
Partial regression plot of the Expanded Disability Status Scale (EDSS) score on immunoglobulin (Ig)G directed against chondroitin-sulfate, p < 0.001.

**Figure 4.**
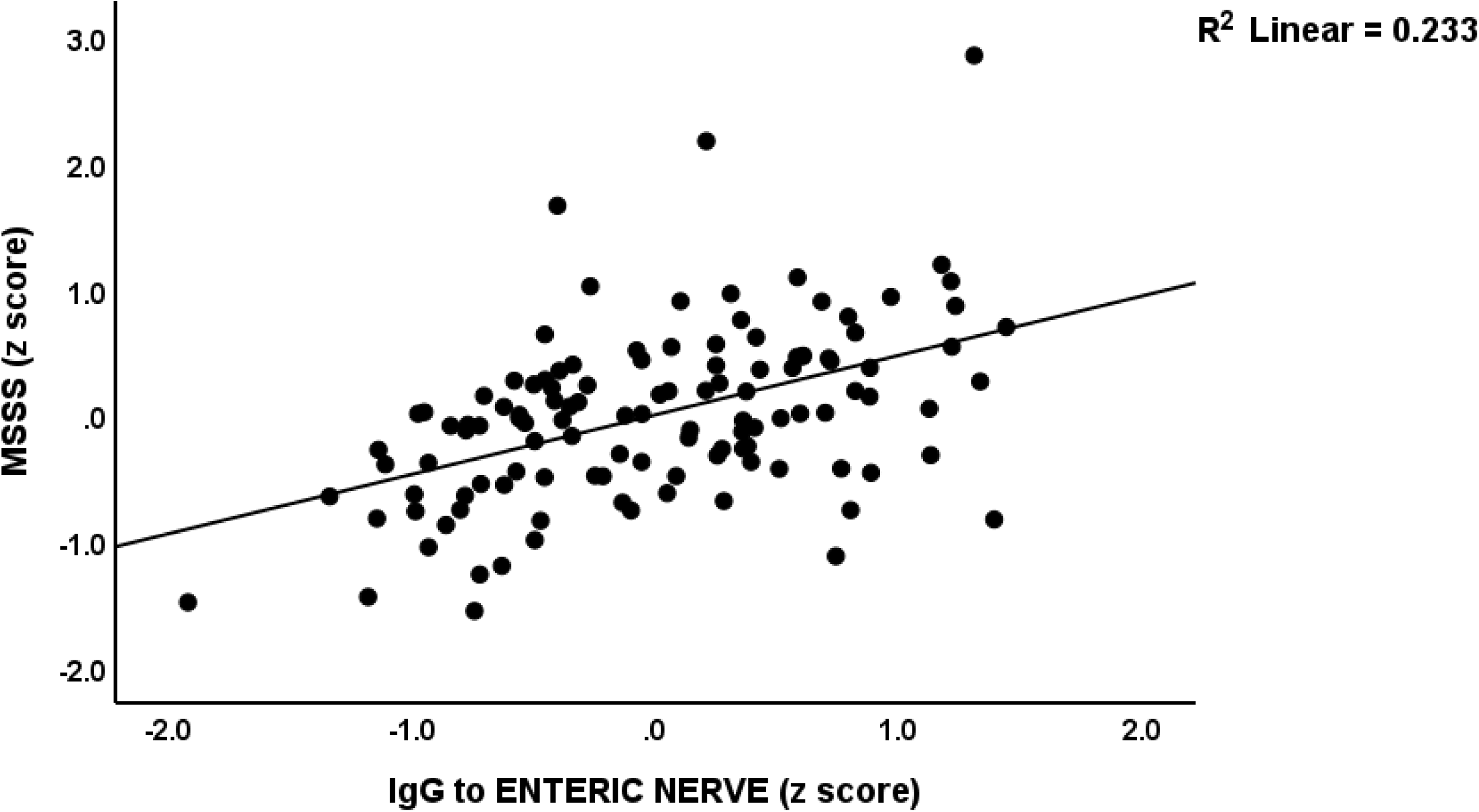
Partial regression plot of the Multiple Sclerosis Severity Scale (MSSS) on immunoglobulin (Ig)G directed against enteric nerve, p < 0.001.

Regression #5 demonstrates that the IRS/CIRS ratio was significantly predicted by IgG-enteric nerve and IgM-Asialo-ganglioside, which together explained 48.7% of its variance. Regression #6 identified IgG-β-crystallin as the exclusive predictor of IRS, accounting for 26.2% of its variability. In this context, CIRS levels were affected by a combination of sex, IgG-enteric nerve, and IgM-synapsin (regression #7), with the model accounting for 61.3% of the total variance.

### Association Between Viral Reactivation and Autoimmune Reactivity to ENS Antigens

To investigate the link between viral antigen exposure and autoimmune responses directed against ENS and non-myelin proteins, we conducted correlation analyses to assess the association between immunoglobulin responses to 9 self-antigens and the PC_3EBNAs and PC_3dUTPases-HHV-6. All IgG/IgA/IgM responses to the self-antigens were significantly correlated with PC_3EBNAs (p<0.001, n=118) and PC_3dUTPases-HHV-6 (p<0.001, n=118).

## The Result of In Silico Analysis

### Common Pentapeptides Among EBV Antigens and ENS

A total of 233 mimicry pentapeptides were identified between EBV antigens and various proteins of the ENS, as presented in ESF2, Table 1. Multiple ENS-related proteins demonstrated significant overlap with specific EBV antigens. As shown in F**igure 5A** Roundabout homolog 1 (ROBO1) exhibited a total of 19 shared pentapeptides, comprising 6 with EBNA1 and EBNA2, 2 with latent membrane protein 2 (LMP2A), 4 with early antigen-D (EA-D), 2 with glycoprotein H (gH), 2 with major capsid protein (MCP), 1 with EBNA6, and 2 with glycoprotein E (gE). RNA-binding protein FUS (Figure 5B) aligned with 15 pentapeptides, including 10 with EBNA1, 2 with LMP1, and 3 with EBNA6.

**Figure 5.**
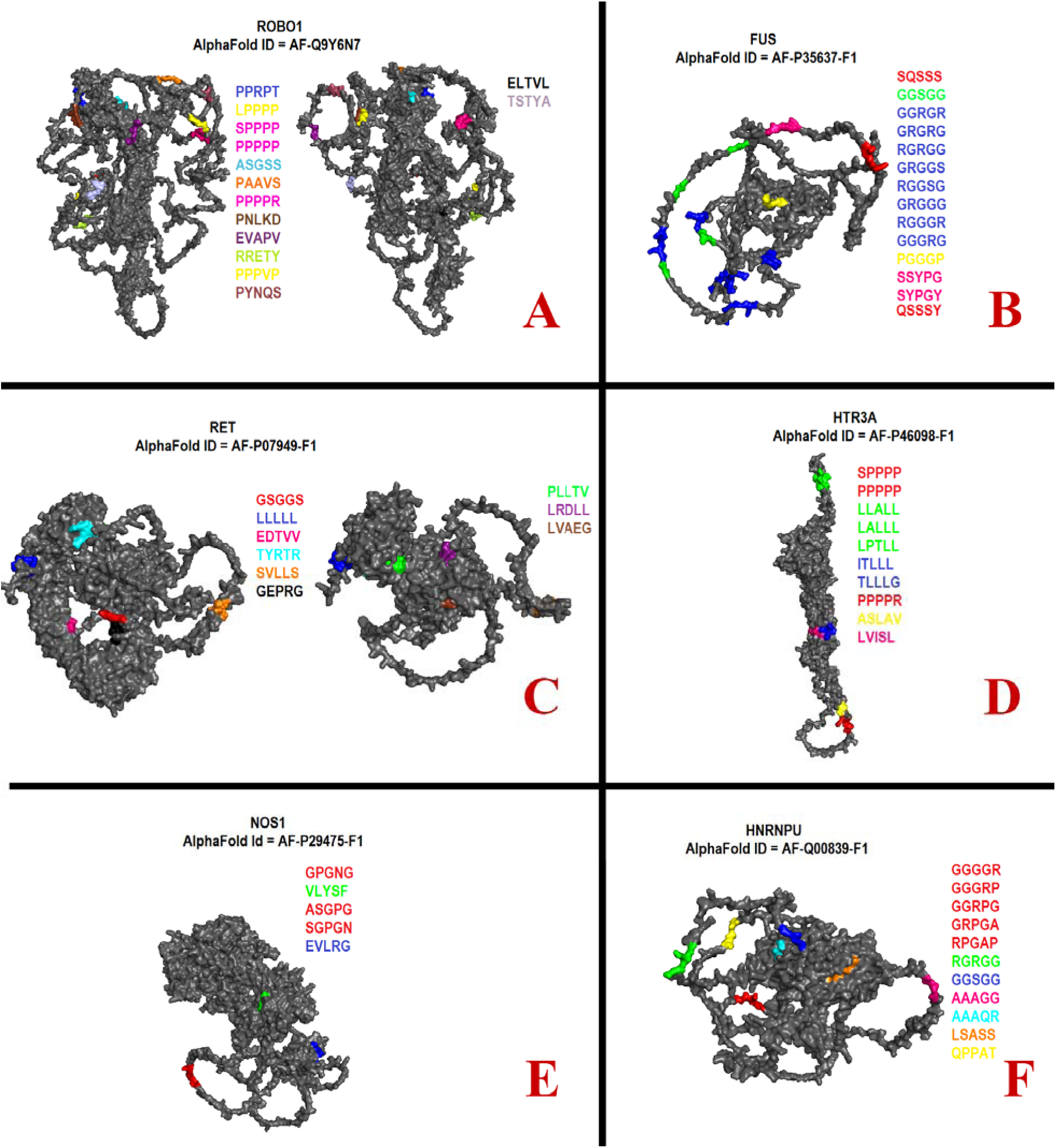
**(A-F)**. Location of Epstein–Barr virus mimicking pentapeptides in 3D structures of Roundabout homolog 1 (ROBO1), RNA-binding protein FUS, proto-oncogene tyrosine-protein kinase receptor RET, 5-hydroxytryptamine receptor 3A (HTR3A), Nitric oxide synthase 1 (NOS1), and Heterogeneous nuclear ribonucleoprotein U (HNRNPU).

As shown in Figure 5C, the proto-oncogene tyrosine-protein kinase receptor RET exhibited 13 shared pentapeptides with EBV, including overlaps with EBNA1, LMP1, LMP2, EA-R, BLLF1, and gB, as well as 3 with gH, 1 with MCP, and 2 with EBNA6. Structural similarity was observed between the 5-hydroxytryptamine receptor 3A (HTR3A, Figure 5D) and EBV antigens, identified through 13 shared pentapeptides: 2 with EBNA2, 5 with LMP1, 1 with LMP2, 3 with EA-D, 1 with BLLF1, and 1 with gB. Nitric oxide synthase 1 (NOS1, Figure 5E) was linked to 12 pentapeptides that are shared with EBV proteins, comprising 2 pentapeptides each with EBNA1 and EBNA2, 1 with LMP1, 2 with BLLF1, 3 with gB, and 1 each with gH and gI. Heterogeneous nuclear ribonucleoprotein U (HNRNPU, Figure 5F) exhibited 12 shared pentapeptides, comprising 7 with EBNA1, 2 with LMP2, 1 with MCP, and 2 with gE. Choline O-acetyltransferase (CHAT) exhibited sequence overlaps across 11 pentapeptides, comprising 3 with EBNA1, 1 with EBNA2, 1 with LMP1, 1 with LMP2, 1 with EA-D, 1 with gB, and 3 with MCP. Further ENS antigens exhibited mimicry, with shared pentapeptides varying from 2 to 9 among different EBV proteins (see ESF2, Table 1).

### Shared Pentapeptides Between HHV-6 Antigens and ENS

The analysis of mimicry patterns between HHV-6 antigens and ENS and other non-myelin proteins identified 311 shared pentapeptides across 36 ENS and non-myelin targets and 28 distinct HHV-6 antigens (see ESF2, Table 2).

As shown in **Figure 6A**, the neurofilament medium polypeptide (NEFM) exhibited the highest degree of molecular mimicry, characterized by 24 shared pentapeptides. The matches comprised gB (2), immediate-early protein 2 (4), U31 (1), U21 (1), U38 (3), dUTPase (1), U51 (2), and U11 (3). Roundabout homolog 1 (ROBO1) exhibited significant overlap, sharing 19 pentapeptides with various HHV-6 antigens (see Figure 6B). The list comprises gB (1), immediate-early protein 2 (3), gH (1), glycoprotein 105 (1), major capsid protein (MCP, 1), dUTPase (1), DNA-binding protein (DBP, 2), U11 (3), and U54 (1).

**Figure 6.**
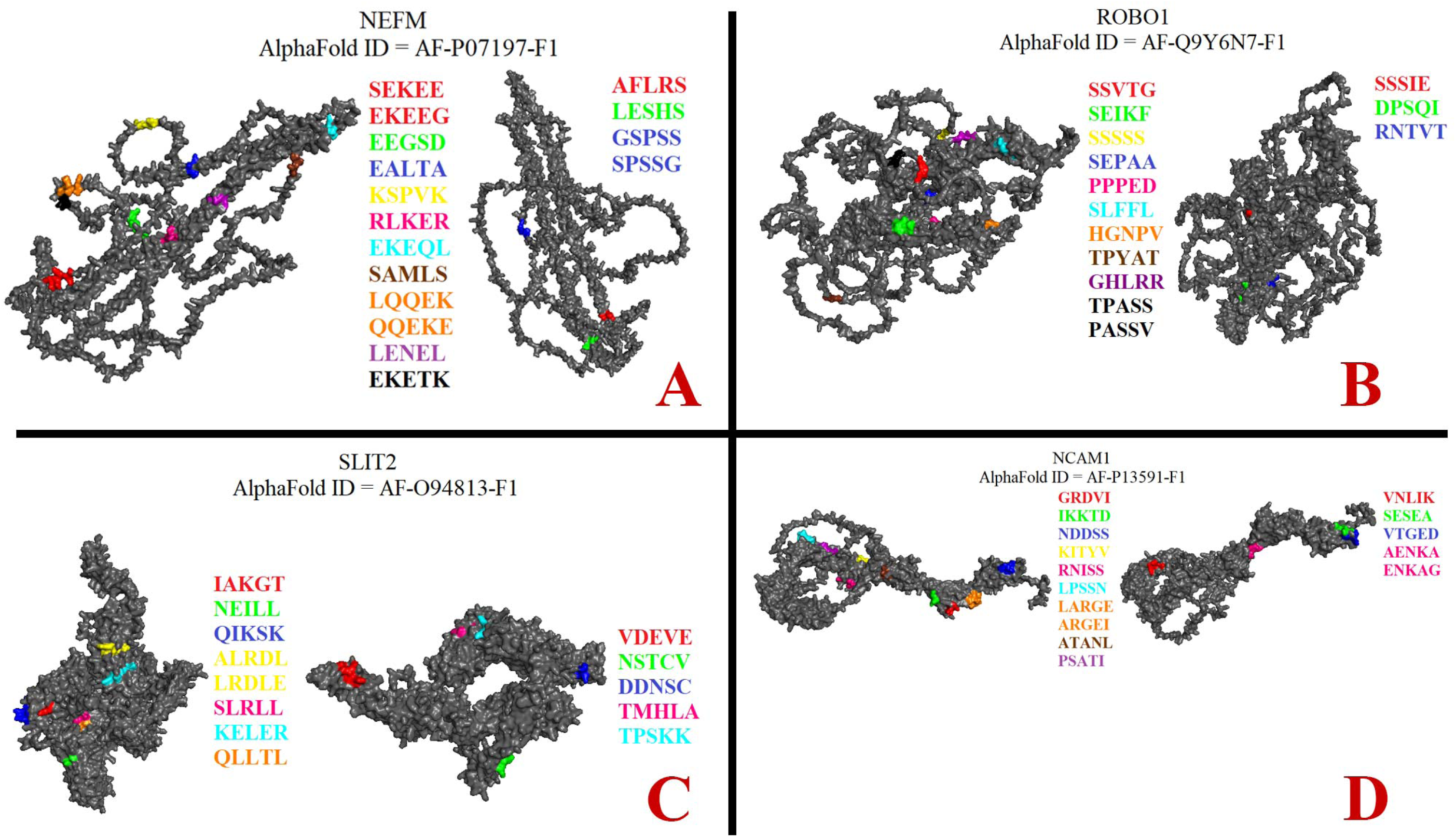
**(A-D)**. Location of human herpesvirus 6 mimicking pentapeptides in 3D structures of neurofilament medium polypeptide (NEFM), Roundabout homolog 1 (ROBO1), SLIT2, neural cell adhesion molecule 1 (NCAM1).

SLIT2 in Figure 6C was linked to 16 shared pentapeptides found in various HHV-6 antigens, including gB (2), immediate-early protein 2 (1), U31 (2), gH (2), glycoprotein 105 (1), U21 (1), RIR (1), U81 (1), U94 (1), and immediate-early protein 1 (1). The neural cell adhesion molecule 1 (NCAM1, Figure 6D) exhibited 16 shared pentapeptides with HHV-6 proteins, such as glycoprotein Q2 (1), U31 (2), gL (1), gH (1), U47 (2), glycoprotein 105 (1), U21 (2), MCP (2), DBP (1), and U94 (3). Other ENS proteins demonstrated significant mimicry. NRG1, RET, L1CAM, and Ku80 exhibited 14 common pentapeptides with different HHV-6 antigens, whereas NEFL presented 13, and both NEFH and NOS1 were linked to 12 shared sequences. Peripherin (PRPH) exhibited 11 mimicry pentapeptides. Other ENS-related proteins, including SNRPA, NTN1, DPP6, AMPH, NOVA1, GDNF, VIP, TUBB3, SOX-10, GAD2, tyrosine 3-monooxygenase, CHAT, P2RX3, HTR3A, CHRNB4, CHRNA3, FUS, TARDBP, TRIM21, SSB, SNRPD1, HTR4, and HNRNPU, demonstrated smaller yet significant degrees of overlap, with each sharing between 2 and 9 pentapeptides with various HHV-6 viral antigens.

### Immunogenic potential of shared pentapeptides

The immunogenic potential of EBV and HHV-6 shared pentapeptides in T and B cell activation is examined and the results are shown in ESF2, Table 3. All of these epitopes (shown in ESF2, Tables 1 and 2) share pentapeptide sequences with self-antigens. Fifty viral antigens containing mimicry pentapeptides were identified as potential T cell activators among EBV-derived epitopes. The most potent T cell immunogens comprised EBNA1, which contained 15 sequences, alongside LMP1 and LMP2, each providing 9 T cell-stimulatory pentapeptides. Other EBV proteins that have shown T cell-activating potential are EBNA2, EA-D, early antigen-R, envelope glycoprotein GP350, gB, gH, and MCP.

In B cell responses, 108 EBV antigens were identified as containing shared pentapeptides that can stimulate antibody production. EBNA1 reemerged as the primary immunogen, containing 43 B cell-reactive pentapeptides. Other significant B cell– activating proteins comprised EA-D, EBNA6, and GP350, each exhibiting 14 reactive sequences; LMP2 with 10; and LMP1 with 9. Other EBV proteins with more restricted B cell activation potential comprised EBNA2 (3 pentapeptides) and early antigen-R (1 pentapeptide). However, none of the mimicry pentapeptides produced from HHV-6 antigens were shown to have the ability to activate B cells, and only two were projected to engage T cells.

## Discussion

### Autoantibodies targeting ENS and CNS antigens in RRMS

The first major finding of this study is that RRMS is characterized by elevated levels of all IgA, IgM, and IgG autoantibodies directed against ENS and other non-myelin antigens (except IgA-Asialo-ganglioside and IgM-synapsin). Most importantly, combinations of these autoantibodies yielded accuracies of 93.7-97.3% in independent holdout samples. IgG targeting the enteric nerve and β-crystallin were the most important predictors.

Prior studies have mainly concentrated on autoantibodies restricted to the CNS, specifically those targeting MBP, MOG, MAG, and GlialCAM [7, 47–50]. Our findings are consistent with the preclinical research by Wunsch et al., which demonstrated that antibody-mediated degeneration of the ENS precedes the development of CNS lesions and the onset of neurological symptoms in experimental models of MS [11]. The degeneration was associated with gastrointestinal dysmotility, enteric gliosis, and neuronal loss.

The autoimmune processes delineated in our study might in theory lead to several peripheral and enteric symptoms extending beyond the CNS. Immune-mediated damage to the enteric nerve might lead to gastrointestinal symptoms, frequently observed in autoimmune diseases, particularly MS [51]. Studies show that autoantibodies targeting the lens protein αβ-crystallin, known for its neuroprotective properties, may contribute to retinal cell death and microglial activation, potentially resulting in cataract formation and neuroinflammatory processes [52] [16, 53]. Antibodies targeting ganglioside such as GM1 are associated with autoimmune peripheral neuropathies, including Guillain-Barré syndrome and chronic autoimmune neuropathies [54, 55]. Peripheral neuropathies, characterized by symptoms such as numbness, tingling, cramping, and muscle twitching, might indicate involvement of the peripheral nervous system [56, 57]. Given the neuroprotective, anti-inflammatory, antioxidant, and antitumor properties of chondroitin sulfate [58, 59], increased autoimmunity might increase neurotoxicity, thereby aggravating disease progression. Moreover, autoimmune-mediated BBB breakdown might facilitate the influx of harmful immune cells and substances leading to demyelination, neurodegeneration, and synaptic dysfunction [60]. Targeting transaldolase may result in a fragment that retains antigenicity, stimulating the proliferation and activity of transaldolase-specific CD8+ T cells in MS patients [61]. Consequently, CD8+ T cells infiltrate the CNS and target oligodendrocytes, leading to their apoptosis and subsequent demyelination [61, 62]. Moreover, the release of perforin and granzymes from these T cells can cause collateral damage to neurons, exacerbating the disease [62].

Autoimmune reactions against other CNS protein also exert harmful consequences, for example autoantibodies targeting GFAP might impair astroglial and lead to white matter loss and BBB instability [63, 64]. While cerebellar autoimmunity causes coordination and cognitive deficits [65, 66], synapsin autoimmunity has been associated with affective symptoms [7].

### Autoimmune profiles and their association with disability and severity

The second major finding of this paper demonstrates a notable relationship between increasing severity of autoimmune responses and clinical disability and severity. IgG levels directed against enteric nerve, β-crystallin, GAFP, asialogangliosides, synapsin, and chondroitin sulphate were significant predictors of EDSS and MSSS scores. This study extends prior research linking CNS-specific autoantibodies (e.g., MBP, MOG, MAG) to the severity of RRMS [7] and suggests that autoimmunity directed against non-myelin associated antigens also plays an important role in the severity of RRMS.

Anti-ganglioside antibody titers, especially IgM and IgG against GM1, GM3, and GD1a are often elevated in patients with secondary progressive or malignant MS [67–69]. Clinical disability levels are positively correlated with enhanced GM1 antibodies [70]. Acarín et al. showed that the primary progressive subtype of MS is significantly linked to anti-GD1a antibodies [69]. Furthermore, neurological disability was positively associated with IgM targeting GD2 [15].

Patients exhibiting more severe or active disease states demonstrate heightened immune reactivity to α-crystallin [71]. However, emerging evidence indicates that anti-αB-crystallin antibodies may constitute a component of the physiological immune repertoire, which may restrict their specificity as biomarkers for disease severity [72]. Additional evidence emphasizes the significance of other autoantibodies in severity of MS, including those against PLP, and beta-2-glycoprotein I, which are associated with increased disability and are more common in progressive MS [73, 74].

### Autoimmune profiles and their association with immune activation

This study identified significant correlations among IgG-enteric nerve, β-crystallin, and GAFP, and IgM and IgA directed against synapsin and Asialo-ganglioside, and immune profiles including the IRS/CIRS ratio, IRS, and CIRS. This indicates that autoimmunity directed to various ENS/CNS self-antigens can augment systemic immune responses thereby promoting disease progression. Almulla et al. found that markers associated with M1 macrophage, Th1, Th2, and Th17, as well as IRS, and CIRS profiles were significantly elevated in RRMS [37]. Kallaur et al. (2017) found that increased levels of interleukin (IL)-10, tumor necrosis factor-α, and interferon-γ coupled with reduced IL-4, were positively correlated with EDSS over a five-year period, underscoring the significance of systemic immune activation in long-term disability [75].

### Viral reactivation and autoimmune targeting of the ENS and CNS

The fourth principal finding highlights the importance of latent viral reactivation in the onset or maintenance of autoimmune responses in RRMS. Significant correlations were identified between EBV and HHV-6 viral markers, particularly EBNA and dUTPases, in conjunction with increased IgA, IgM, and IgG responses to non-myelin-associated self-antigens, including ENS, crystallin, BBB, ganglioside, transaldolase and cerebellar self-antigens. Jakimovski et al. demonstrated a correlation between elevated EBV titers and grey matter loss, as well as increased severity of MRI-detected pathology in RRMS [76]. Prior studies demonstrate that viral reactivation induces pro-inflammatory cytokines and atypical T-cell responses, which play a role in neurodegeneration [29, 77]. Liu et al. reported that viral infections, particularly EBV in the CNS, may elevate the risk of GFAP autoimmunity development [78].

### Viral antigens mimic ENS proteins

In-silico analysis found a substantial pentapeptide overlap among 41 viral proteins (13 from EBV and 28 from HHV-6) and 36 human proteins linked to the enteric nerve. This study extends prior research on mimicry associated with CNS proteins [31, 50] and provides new evidence suggesting that mimicry is relevant to peripheral targets as well. Previous studies indicate that the immunodominant epitope of human transaldolase (residues 271-285) exhibits cross-reactivity with peptides from EBV and HSV type 1, suggesting that molecular mimicry may trigger or exacerbate autoimmunity in MS [17, 18]

Our study shows that many identified viral–ENS peptide matches display predicted immunogenicity for T-cells and B-cells. Two pentapeptides derived from HHV-6 exhibited T-cell immunogenicity, whereas none displayed immunogenicity to B cells, consistent with prior research on latent viral and CNS protein mimicry [31]. Mimicry events may operate via MHC-II-mediated antigen presentation, resulting in the activation of autoreactive CD4+ T cells [79, 80]. EBV and HHV-6 are acknowledged for their involvement in the proliferation of autoreactive T and B cell populations and the induction of cytokines like IFN-γ and TNF-α, which contribute to neuroinflammation [81, 82].

## Limitations

In this present study, several limitations should be considered while interpreting the findings. First, the connection between GI symptoms and ENS-targeted autoantibodies has not been formally evaluated using validated clinical GI symptom scales. While existing literature suggests a connection [11] our current study does not conclusively demonstrate an association between serological findings and functional enteric dysfunction in patients with RRMS. Second, this study solely focused on RRMS. Future studies should explore these autoimmune biomarkers in other subtypes of MS including secondary progressive MS.

Third, in-silico findings are predictive and lack experimental validation. While pentapeptide homology and immunogenicity scores indicate potential T-cell activation, direct validation via T-cell reactivity assays or epitope mapping was not conducted. Finally, viral reactivation was deduced from serological markers (IgA, IgM, and IgG against EBNA and dUTPases), instead of through direct measurement of viral DNA or RNA in blood or tissue samples. Immunoglobulin profiles indicate immune exposure and reactivation; however, additional research employing PCR-based or next-generation sequencing methods is necessary to validate viral persistence or replication.

## Conclusions

This study’s findings reveal elevated levels of autoantibodies directed against antigens in both the ENS and CNS in RRMS. Those autoimmune responses demonstrated a notable correlation with disability, severity, and systemic immune activation. The reactivation of EBV and HHV-6 correlated with the presence of these autoantibodies, and in-silico analyses supported a molecular mimicry mechanism. The findings highlight the role of the enteric nerve and self, non-myelin antigens as clinically relevant, though previously underrecognized, factors in the pathophysiology of RRMS.

## Data Availability

The corresponding author (MM) will grant access to the dataset supporting this study upon receipt of a valid request and the completion of a comprehensive data review.

## Acknowledgements

The authors express sincere gratitude to the Neuroscience Centre of Alsader Medical City in Al-Najaf province, Iraq, for their significant assistance in the data collection process.

## Ethical approval and consent to participate

The Ethics Committee of the College of Medical Technology at the Islamic University of Najaf, Iraq, granted sanction for the investigation (Document No. 11/2021). Written informed consent was obtained from all patients and control participants, and all procedures were conducted in accordance with Iraqi and international ethical standards.

## Author’s contributions

AFA oversaw the collection of blood samples and the procedures involving patients. Quantification of serum biomarkers was conducted by AV at Immunosciences Lab, which covered the testing expenses. MM performed the statistical analysis. MM and AV developed visual representations. MGN performed an analysis of molecular mimicry. The initial manuscript was drafted by AFA and subsequently reviewed and revised by MM, AV, EV, and YZ. All authors consented to the final version of the manuscript.

