## supplementary file 1 for "Autoantibodies targeting the enteric nerve and non-myelin epitopes in relapsing-remitting multiple sclerosis: diagnostic relevance and viral mimicry"

**ELECTRONIC SUPPLEMENTARY FILE (ESF)**

#: Joined first authorships

(1) Sichuan Provincial Center for Mental Health, Sichuan Provincial People's Hospital, School of Medicine, University of Electronic Science and Technology of China, Chengdu 610072, China.
(2) Key Laboratory of Psychosomatic Medicine, Chinese Academy of Medical Sciences, Chengdu 610072, China.
(3) Medical Laboratory Technology Department, College of Medical Technology, The Islamic University, Najaf, 31001, Iraq.
(4) Department of Pathology, Faculty of Medicine, Saint Petersburg State University, 199034 Saint Petersburg, Russia.
(5) Department of Psychiatry, Faculty of Medicine, Chulalongkorn University, Bangkok, Thailand.
(6) Cognitive Fitness and Technology Research Unit, Faculty of Medicine, Chulalongkorn University, Bangkok, Thailand.
(7) Department of Psychiatry, Medical University of Plovdiv, Plovdiv, Bulgaria.
(8) Research Institute, Medical University Plovdiv, Plovdiv, Bulgaria.

Kyung Hee University, 26 Kyungheedae-ro, Dongdaemun-gu, Seoul 02447, Republic of Korea.

**ESF, Table 1.** Cytokine, chemokines and growth factors examined in the current study

| **Protein abbreviations** | Gene Symbol | **Protein name** |
| --- | --- | --- |
| IL-1β | IL1B | Interleukin-1β |
| IL-1RA | IL1RN | Interleukin-1 receptor antagonist |
| IL-2 | IL2 | Interleukin-2 |
| IL-4 | IL4 | Interleukin-4 |
| IL-5 | IL5 | Interleukin-5 |
| IL-6 | IL6 | Interleukin-6 |
| IL-7 | IL7 | Interleukin-7 |
| CXCL8 | CXCL8 | C-X-C motif chemokine ligand 8 (IL-8) |
| IL-9 | IL9 | Interleukin-9 |
| IL-10 | IL10 | Interleukin-10 |
| IL-12p70 | IL12 | Interleukin-12 |
| IL-13 | IL13 | Interleukin-13 |
| IL-15 | IL15 | Interleukin-15 |
| IL-17 | IL17 | Interleukin-17 |
| CCL11 | CCL11 | Eotaxin |
| FGF2 | FGF2 | Fibroblast growth factor 2, Basic fibroblast growth factor |
| G-CSF | CSF3 | Granulocyte Colony Stimulating Factor, Colony Stimulating Factor 3 (Granulocyte) |
| GM-CSF | CSF2 | Granulocyte-macrophage colony-stimulating factor, Colony-stimulating factor 2 |
| IFN-γ | IFNG | Interferon-γ |
| CXCL10 | CXCL10 | C-X-C motif chemokine ligand 8, Interferon gamma-induced protein 10 (IP10) |
| CCL2 | CCL2 | C-C Motif Chemokine Ligand 2 (MCP1) |
| CCL3 / MIP-1α | CCL3 | Macrophage inflammatory protein-1 alpha, C-C Motif Chemokine Ligand 3 |
| PDGF | PDGFA | Platelet Derived Growth Factor Subunit A |
| CCL4 / MIP-1β | CCL4 | C-C Motif Chemokine Ligand 4, Macrophage Inflammatory Protein 1-Beta, Lymphocyte Activation Gene 1 Protein |
| CCL5 /RANTES | CCL5 | C-C Motif Chemokine Ligand 5, Regulated Upon Activation, Normally T-Expressed, And Presumably Secreted |
| TNF-α | TNF | Tumor Necrosis Factor-Alpha |
| VEGF | VEGFA | Vascular Endothelial Growth Factor |

**ESF, Table 2**. Description of the immune profiles used in this study

| **Immune Profile** | **Members** |
| --- | --- |
| **IRS** | IL-1β, IL-6, TNF-α, CXCL8, CCL3, IL-2, IFN-γ, IL-12, IL-17, IL-15, G-CSF, GM-CSF, CXCL10, CCL5, CCL2 |
| **CIRS** | IL-4, IL-10, sIL-1RA |
| **IRS+CIRS** | IL-1β, IL-6, TNF-α, CXCL8, CCL3, IL-2, IFN-γ, IL-12, IL-17, IL-15, G-CSF, GM-CSF, CXCL10, CCL5, CCL2, IL-4, IL-10, sIL-1RA |

IRS: immune-inflammatory response system; CIRS: compensatory immunoregulatory system

**Table 3.** Sociodemographic and clinical data in patients with relapsing-remitting multiple sclerosis (RRMS) and healthy controls (HC).

| **Variables** | **Healthy Controls (n=63)** | **RRMS Patients (n=55)** | **F/χ^2^** | **df** | **p-value** |
| --- | --- | --- | --- | --- | --- |
| Age (years) | 31.4 (7.1) | 29.5 (6.3) | 2.18 | 1/116 | 0.142 |
| Sex (M/F) | 33/30 | 37/18 | 2.70 | 1 | 0.133 |
| BMI (kg/m2) | 26.33 (4.50) | 24.94 (3.52) | 3.41 | 1/116 | 0.068 |
| Education (years) | 17.2 (5.1) | 18.4 (3.4) | 2.04 | 1/116 | 0.155 |
| Married/Separated | 26/37 | 31/24 | 2.68 | 1 | 0.139 |
| Smoking (N/Y) | 44/19 | 50/5 | 8.04 | 1 | 0.006 |
| Employment (N/Y) | 2/28 | 46/9 | 46.78 | 1 | <0.001 |
| MSSS | 0 | 1.71 (1.20) | MWU | - | <0.001 |
| EDSS | 0 | 1.03 (0.11) | MWU | - | <0.001 |
| PC-Severity | -0.921(0) | 1.05(0.198) | MWU | 1/116 | <0.001 |
| PC_3dUTPase-EBV | -0.662(0.745) | 0.759(0.655) | 119.51 | 1/116 | <0.001 |
| PC_3dUTPase-HHV-6 | -0.697(0.694) | 0.798(0.628) | 148.71 | 1/116 | <0.001 |
| PC_3EBNA | -0.579(0.810) | 0.664(0.755) | 73.71 | 1/116 | <0.001 |

M: Male, F: Female, kg: Kilogram, m2: squared meter, N: No, Y: Yes, MSSS: Multiple Sclerosis Severity Score, EDSS: Expanded Disability Status Scale, PC-Severity: first principal component extracted from EDSS and MSSS, PC 3dUTPase-EBV: first principal component extracted from IgA, IgM,IgG directed to Epstein-Barr Virus deoxyuridine-triphosphatase (dUTPase-EBV), PC 3dUTPase-HHV-6: first principal component extracted from IgA, IgM,IgG directed to herpesvirus 6 deoxyuridine-triphosphatase (dUTPase-HHV-6), PC 6dUTPases: first principal component extracted from IgA, IgM,IgG directed to both dUTPase-EBV and dUTPase-HHV-6, PC 3EBNA: first principal component extracted from IgA, IgM,IgG directed to Epstein–Barr virus nuclear antigen 1 (EBNA).
